# Proteomic profiling of postmortem prefrontal cortex identifies molecular signatures of borderline personality disorder

**DOI:** 10.64898/2026.11.09.26367584

**Authors:** Eric Zillich, Diana Avetyan, Christian C. Witt, Lea Zillich, Josef Frank, Frank Stein, Jennifer J. Schwarz, Naguib Mechawar, Gustavo Turecki, Rainer Spanagel, Fabian Streit, Stephanie H. Witt

## Abstract

Borderline personality disorder (BPD) is characterized by a severe pattern of symptoms including emotional instability, impulsivity, and unstable interpersonal relationships. A complex interplay between genetics and environmental factors has been hypothesized to be associated with the development of BPD. While neuroimaging studies have identified structural and functional changes in the brain, and genome-wide association studies described some first genetic risk loci, the neurobiological underpinnings of BPD remain poorly described. To better understand disease processes in BPD, we performed a molecular characterization of the prefrontal cortex (PFC), an important brain region for emotion and impulsivity regulation in BPD. Label-free brain proteomics was performed in postmortem human brain tissue in a sample of n=79, with a case group of n=36 individuals with BPD and a control group of n=43 individuals without BPD. Differential protein expression analysis suggested protein-level dysregulation of mitochondrial, immunological, and neuroplasticity pathways. Network analyses identified a BPD-associated co-expression module enriched for protein secretory and synaptic processes that is druggable with selective serotonin reuptake inhibitors. Additional druggability analyses on protein-protein interaction networks and drug-repositioning analysis suggested VEGF signaling as a potential pharmacologic target in BPD. When we integrated BPD proteomic signatures with genomic and proteomic datasets of common comorbidities such as posttraumatic stress disorder and major depressive disorder, we observed a strong convergent overlap across disorders, with STX7 and RTN4 identified as conserved, differentially expressed proteins. Our study provides first insights into proteomic signatures of BPD in the brain, suggests potential pharmacological targets, and provides evidence for convergent protein signatures across BPD and common comorbidites.

## Background

Borderline personality disorder (BPD) is a severe mental disorder affecting around 1% of the general population, while prevalence estimates range up to 20% in the psychiatric inpatient population(1). BPD is characterized by a set of key symptoms including impulsivity, emotional instability, an unstable self-image, and difficulties in interpersonal relationships(2). Further, strong patterns of self-harm and suicidal ideation as well as suicide attempts are observed in individuals with BPD(3). The typical onset of BPD is in adolescence and individuals with BPD often suffer for years until treatment is initiated. Even with psychotherapy as the gold-standard treatment, a substantial fraction of patients remain with severe BPD symptoms that negatively impact their daily lives(4). The clinical burden of BPD is further exacerbated by high rates of somatic as well as psychiatric disorder co-diagnoses(5–7) resulting in a reduced life expectancy of individuals with BPD of 6-32 years compared to the general population(3, 8).

The etiology of BPD is complex, and genetic as well as environmental factors contributing to BPD risk have been identified(9). The heritability of BPD has been estimated at ∼46% in twin studies(10) and a recent genome-wide association study (GWAS) has identified 11 genome-wide significant risk loci for BPD, involving genes such as *FOXP2, EXD3, and SGCD*(*11*). Among the most important environmental factors are adverse childhood experiences (ACE)(12, 13). Individuals with BPD are around 14 times more likely to have experienced childhood adversity compared to non-clinical cohorts(12). Exposure to early life stress and traumatic experiences was associated with hypothalamic-pituitary-adrenal (HPA) axis dysregulation(14) and DNA methylation changes in BPD(15), thereby establishing a link between genetic and environmental risk factors.

In terms of neural correlates of BPD, a fronto-limbic dysbalance has been proposed, characterized by hyperactivity of limbic areas such as amygdala and hippocampus and an impaired top-down control due to frontal cortical hypoactivity(16, 17). While this model could explain increased impulsivity and impaired emotional regulation in BPD, other studies have pointed to a lack of brain region specificity and suggested strong context-dependence of neuroimaging findings(18, 19). Knowledge about molecular correlates of BPD is limited and reseach mainly focused on candidate genes including *NR3C1*, *BDNF*, and *IL6*, where epigenetic(20) and transcriptional changes(21) were investigated. Some early epigenome-wide association studies of BPD have been performed in blood, identifying differentially methylated CpG sites in BPD; however, results strongly varied across studies(22–24).

Studies investigating molecular signatures of psychiatric disorders in the brain are scarce due to limited availability of postmortem brain tissue, and to our knowledge, have not been performed in BPD so far. Further, in contrast to epigenetic and transcriptional changes that have at least been reported in blood for BPD(21–24), changes at the protein level remain poorly studied. One recent study investigated the hair proteome in BPD and identified a predictive model that was able to distinguish patient and control groups based on proteomic data(25). Molecular functions related to the predictive model were related to translation, oxidative phosphorylation, and oxidative stress pathways.

For a deeper understanding of the neurobiology of BPD, we aimed to identify protein and protein network changes in the brain associated with BPD by comparing the proteome of individuals with BPD with an unaffected control group in postmortem brain tissue. We focused our proteomic study on the prefrontal cortex (PFC) due to its important role in the regulation of emotions, aggression, and impulsivity in BPD(26, 27). The investigated Brodmann Area 10 (BA10), known as the frontopolar PFC, also has a particular role in pain processing, which is known to be impaired in BPD(28, 29). A characterization of biological processes affected by proteomic changes in BPD was performed using pathway enrichment analyses and by constructing protein-protein interaction networks. In addition, we aimed to evaluate potential novel pharmacotherapeutic options in BPD by performing drug repositioning analyses and integrated our findings with genomic and proteomic datasets of other psychiatric disorders associated with BPD.

## Methods

### Human postmortem brain tissue

Postmortem human brain tissue of the prefrontal cortex (PFC, n=79) was obtained from the Douglas Brain Bank (DBB) in Montréal, Canada. Tissue samples were taken from BA10 which is located at the frontal brain pole and is also known as the frontopolar PFC. Phenotypic profiling of sample donors at DBB includes a standardized psychological autopsy, including next-of-kin interviews, with additional information obtained from coroner’s notes. For our case-control study of proteomic signatures of BPD (n=36 BPD, n=43 Ctrl), during sample selection, we aimed for a balanced distribution of covariates such as age, sex, PMI, pH, and comorbid psychiatric disorders such as substance use disorder (SUD) and major depressive disorder (MDD) across groups. Inclusion criteria were age > 18 and BPD diagnosis status (yes/no) according to DSM-IV, based on a SCID-II interview. Donors with schizophrenia (SCZ) or bipolar disorder (BIP) subtype I diagnoses were excluded from the study. To minimize confounding effects of ethnic background on proteomic signatures, we aimed for a homogeneous cohort and analyzed the ethnic group with the largest number of available samples at DBB, which was European American (EA), resulting in a final selection of n=79 PFC samples.

### Protein extraction

Fresh-frozen PFC postmortem brain tissue was stored at -80°C until protein extraction. Samples were randomized based on BPD status and covariates during probe handling. For tissue lysis, RIPA buffer (Pierce, Thermo Fischer) with cOmplete^TM^ Mini Protease inhibitor (Roche) was prepared and stored on ice. A total volume of 300µl lysis buffer was transferred to a 1.5ml Eppendorf tube, and a 30mg piece of postmortem brain tissue was added. Using a plastic pestle (Bel-Art®), the tissue was thoroughly dissociated, followed by a 30min lysis period on ice. Every 5 minutes, the lysate was homogenized using a vortex mixer to facilitate tissue lysis. The lysate was then centrifuged at 10.000g for 5 minutes at 4°C. The supernatant, corresponding to the protein lysate of PFC samples, was transferred to another sample tube. Protein concetration was measured using Direct Detect® Assay-free membrane cards and a Direct Detect® Spectrometer system (both Merck Millipore). Samples were randomized on a 96-well plate with 20µg of total protein diluted in 60µl protease inhibitor containing RIPA buffer. The plate was stored at -80°C until proteomic analysis.

### Mass spectrometry

An UltiMate 3000 RSLCnano LC system (Thermo Fisher Scientific) equipped with a trapping cartridge (µ-Precolumn C18 PepMap™ 100, 300 µm i.d. × 5 mm, 5 µm particle size, 100 Å pore size; Thermo Fisher Scientific) and an analytical column (nanoEase™ M/Z HSS T3, 75 µm i.d. × 250 mm, 1.8 µm particle size, 100 Å pore size; Waters) was used. Samples were trapped at a constant flow rate of 30 µL/min using 0.05% trifluoroacetic acid (TFA) in water for 6 minutes. After switching in-line with the analytical column, which was pre-equilibrated with solvent A (3% dimethyl sulfoxide [DMSO], 0.1% formic acid in water), the peptides were eluted at a constant flow rate of 0.3 µL/min using a gradient of increasing solvent B concentration (3% DMSO, 0.1% formic acid in acetonitrile).

The outlet of the analytical column was directly coupled to an Orbitrap Exploris™ 480 mass spectrometer (Thermo Fisher Scientific) equipped with a Nanospray Flex™ ion source, operating in positive ion mode. Peptides were introduced into the instrument using a Pico-Tip emitter (360 µm OD × 20 µm ID; 10 µm tip, CoAnn Technologies) with a spray voltage of 2.5 kV. The capillary temperature was maintained at 275 °C. Full MS (MS1) scans were acquired in centroid mode over an m/z range of 420–680, with a resolution of 60,000 at m/z 200 in the Orbitrap. The automatic gain control (AGC) target was set to ‘custom’ with a normalized AGC target of 300%. Data-independent acquisition (DIA) was performed across a precursor mass range of 430–670 m/z, using 4 m/z isolation windows. MS/MS scans were acquired in centroid mode over a scan range of 200–1800 m/z, with a resolution of 30,000. The AGC target for DIA was set to ‘custom’, with a normalized value of 3000%. A normalized higher-energy collisional dissociation (HCD) energy of 28% was applied for fragmentation.

### Proteomics data preprocessing

Preprocessing and analysis of proteomic data were performed based on a modified version of an analysis script by Frank Stein (EMBL, Heidelberg, Germany). All preprocessing and analysis steps were conducted in R 4.4.0(30).

Raw files were analyzed using DIA-NN 1.8.1(31) with directDIA module using an in silico DIA-NN predicted spectral library (Trypsin as protease; C carbamidomethylation and N-terminal M excision; Protein N-terminal acetylation and Oxidation on Methionine enabled; 2 missed cleavages; precursor m/z range: 430-670; precursor charge range 2 to 4). The spectral library was generated from fasta files downloaded from Uniprot (homo sapiens, UP000005640, ID9606, 20,594 entries, October 2022). The DIA-NN search used the following parameters: Precursor FDR (%)= 1; Mass accuracy, MS1 accuracy, and Scan window = 0; Use isotopologues; match between runs enabled; No shared spectra; Protein inference: Genes, Neural network classifier: Single-pass mode; Quantification strategy: Robust LC (high precision); Cross-run normalization: RT-dependent; Library generation: Smart profiling; Speed and RAM usage: optimal results.

DIA-NN output (report.tsv files) was processed to generate filtered results at the protein level for further downstream analysis. To ensure high data quality, the following filtering criteria were applied to the DIA-NN results: Q.Value <= 0.01, PG.Q.Value <= 0.01, and Lib.PG.Q.Value <= 0.01. For proteins lacking gene annotations, the ‘Protein.ID’ was used to fill in missing ‘Gene’ entries, ensuring comprehensive gene-level representation. The filtered tables ‘diann_outputs_filtered_report.uniqueGenes_maxLFQ_matrix.tsv’ and ‘diann_outputs_filtered_report.ProteinGroups_maxLFQ_matrix.tsv’ were generated based on the ‘Genes.MaxLFQ.Unique’ and ‘PG.MaxLFQ’ columns of the DiaNN report output, respectively. These matrices include protein-level data, enriched with protein-level summaries, such as the number of Razor Peptides, Unique Peptides, Total Peptides, and the Total Intensity, which were calculated on the combined dataset and integrated into the gene-level output. The quantification is based on all peptides.

For proteomic data analysis, initial data processing included filtering out contaminants and reverse proteins. Only proteins quantified with at least 2 razor peptides (with Razor.Peptides >= 2) were considered for further analysis. Additionally, only proteins identified and quantified in at least two thirds (2/3) of the number of replicates from each condition were retained to ensure robustness. 6819 proteins passed the quality control filters. Log2 transformed raw DIA intensities values (’Intensity’ columns) were normalized using the ‘normalizeQuantiles’ function of the limma package. Missing values were imputed with the ‘knn’ method using the ‘impute’ function from the Msnbase package(32). This method estimates missing data points based on similarity to neighboring data points, ensuring that incomplete data did not distort the analysis.

### Differential protein expression analysis

Differential protein expression analysis on the imputed dataset was performed for BPD case/control status using the moderated t-test provided by the limma package(33). The model accounted for covariates including age, sex, PMI, pH, SUD and MDD status in the design matrix passed to the ‘lmFit’ function. Imputed values were assigned a weight of 0.01 in the model, while quantified values were given a weight of 1, ensuring that the statistical analysis reflected the uncertainty in imputed data. Resulting p-values were FDR-corrected to adjust for multiple testing. Differential expression criteria for proteins were FDR q<0.05 for proteome-wide significance and p<0.05 for nominal significance.

### Gene set enrichment analysis

To better understand the biological relevance of differential protein expression signatures in BPD, we performed gene set enrichment analysis (GSEA) on limma results. Results were ranked by the limma t-statistic, and the gseGO function of clusterProfiler(34) was used for GSEA based on Gene Ontology categories: biological process (BP), molecular function (MF), and cellular component (CC). GSEA results with FDR q<0.05 were considered statistically significant and were visualized as enrichment map (emap) plots(35).

### Weighted gene co-expression network analysis

To complement the differential protein expression analysis, focused on identifying single protein associations with BPD, we performed a protein network-centered approach by applying weighted gene co-expression network analysis (WGCNA)(36) on the proteomic dataset. The normalized dataset without imputation of missing values was used for WGCNA to ensure that protein co-expression networks are derived from measured intensities only. During network construction, soft threshold picking was performed based on R^2^>0.85 and mean connectivity < 100 criteria, resulting in a soft threshold of 5. The blockwiseModules function was applied with parameters TOMType=”signed”, minModuleSize=20, and mergeCutHeight=0.1. Protein co-expression networks were associated with phenotypic variables to identify potential BPD-associated modules by calculating Pearson correlation coefficients with the module eigengene. A GO enrichment analysis was performed on module genes to characterize the biological function of protein networks. Overrepresentation of network genes within GO BP, MF, and CC pathways with FDR q<0.05 was considered statistically significant.

### Protein-protein interaction (PPI) networks

We constructed protein-protein interaction (PPI) networks to further investigate the role of BPD-associated proteins in biological networks. The STRING database 12.0 (https://string-db.org)(37) was used to construct PPI networks for i) nominal significant DEPs and ii) WGCNA module lightyellow proteins. Networks were restricted to contain only high-confidence protein-protein interactions (>0.7), and singletons were excluded. Downstream analyses of the PPI networks included the identification of network hub genes defined as highly connected nodes, as well as a pathway enrichment analysis to investigate the biological processes related to the PPI networks.

### Drug repositioning analysis

Drug repositioning analysis was performed on molecular signatures of the NIH LINCS L1000 dataset(38) as implemented in the CMap query tool (https://clue.io/query). As the query dataset, we used the top 150 upregulated and downregulated proteins from differential protein expression analysis, sorted by the limma t-statistic. In CMap, connectivity scores are calculated between the query and the reference signature. Reference signatures include gene expression signatures from drug screening and gene overexpression/knockdown studies. Results of the drug repositioning analysis are reported at the level of individual perturbagens i.e. drugs or genes, but also at the level of pathways and mechanisms of action. We complemented the signature-based analysis by performing a druggability analysis of BPD-associated WGCNA network proteins and PPI network proteins using DGIdb(39).

### GWAS enrichment analysis

To integrate the brain proteomic dataset with information on genetic risk variants for psychiatric disorders, we performed a GWAS enrichment analysis using MAGMA(40). GWAS summary statistics for BPD as well as for psychiatric disorders with strong co-occurrence with BPD were obtained from the PGC GWAS repository (https://pgc.unc.edu/for-researchers/download-results/). Publicly available GWAS summary statistics from the most recent GWAS in European American ancestry were used for the GWAS enrichment analysis to match the ethnic background of the postmortem brain cohort. Investigated phenotypes in addition to BPD due to high rates of co-diagnoses include attention deficit hyperactivity disorder (ADHD)(41), bipolar disorder (BIP)(42), major depressive disorder (MDD)(43), posttraumatic stress disorder (PTSD)(44), schizophrenia (SCZ)(45), and substance use disorder (SUD)(46). Gene sets for the enrichment analysis in MAGMA were derived from differential protein expression analysis with upregulated and downreguated proteins associated with BPD (nominal p<0.05), as well as BPD-associated module lightyellow proteins from WGCNA. Results of gene set-based tests were adjusted for multiple testing using Bonferroni correction (n=7 tests).

### Integrative analyses with PTSD and MDD proteomic datasets

We performed an integrative analysis of BPD-associated proteomic signatures with data from Wang, Liu (47) that investigated postmortem dorsolateral prefrontal cortex proteomic signatures of MDD and PTSD, two diagnoses highly comorbid with BPD. The proteomic dataset from Wang, Liu (47) originated from Brodmann Areas 9 and 46 that are directly adjacent to BA10. First, an overlap analysis of nominally significant differentially expressed proteins from the case-control comparisons was performed using a venn diagram. Next, we performed rank-rank hypergeometric overlap analysis (RRHO) using RRHO2(48) based on full differential expression statistics to identify concordant and discordant protein expression patterns across BPD, MDD, and PTSD.

## Results

We first investigated demographic statistics of the BPD postmortem brain cohort (n=79, Table 1). Among the available phenotypic variables, we found significant group differences for cause of death (p=5.6e-14) and brain pH (p=0.002), while all other variables did not differ between the BPD and control group (p>0.05). We found BPD brains to have a significantly higher pH value and in addition, that tissue donors with BPD died significanly more often by suicide compared to control individuals.

**Table 1:**
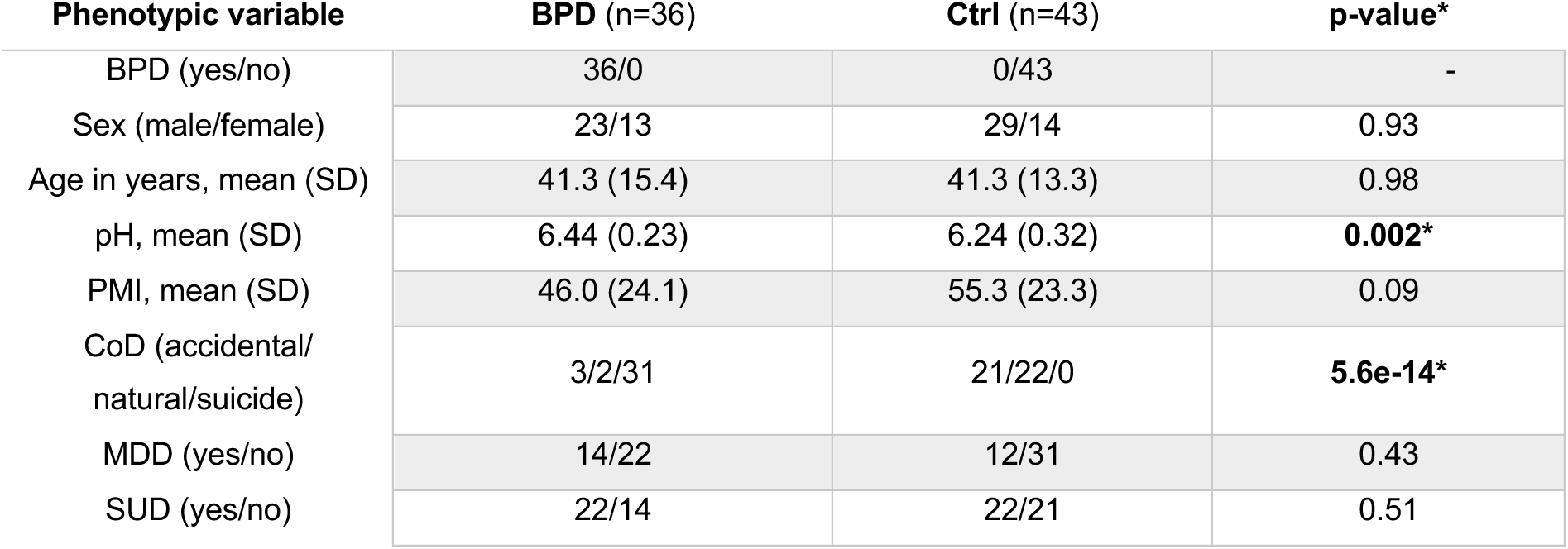
Demographic statistics for the cohort of postmortem brain tissue donors (n=79) included in the proteomic analysis. Group differences were investigated using t-tests and χ^2^ test for continuous and discrete phenotypic variables, respectively. BPD: borderline personality disorder, Ctrl: control group, SD: standard deviation, pH: postmortem brain tissue pH, PMI: postmortem interval, CoD: cause of death, MDD: major depressive disorder, SUD: substance use disorder.

Proteomic profiling of PFC postmortem brain samples was performed using label-free mass spectrometry in data-independent acquisiton (DIA) mode (Figure 1A). The number of identified proteins per sample passing QC filtering ranged between 6,226 and 6,900 (Figure S1A). Raw data was vsn-normalized and k-nearest-neighbour-based imputation was performed to reduce missigness in the dataset (Figure S1A, see Methods section for details). To identify proteomic signatures of BPD in the PFC, we performed a differential protein expression analysis for the final set of 6,819 proteins between BPD and Ctrl samples in limma while adjusting for covariates age, sex, PMI, pH, SUD and MDD status (Table S1). We identified a total of 229 differentially expressed proteins (DEPs) at nominal significance (p<0.05) of which 100 were upregulated (log2FC>0) and 129 were downregulated (log2FC < 0), while no association remained signifinant after multiple testing correction (q<0.05, Figure 1B). The protein *cytosolic arginine sensor for mTORC1 subunit 1* (CASTOR1, log2FC=-0.25, p=0.001, q=1) showed the strongest downregulation in BPD, while *Microtubule Associated Protein 9* (MAP9, log2FC=0.13, p=0.003, q=1) was the top upregulated protein. To investigate the biological role of differential protein expression signatures in BPD, we performed a gene set enrichment analysis (GSEA). Among significant results, positive GSEA normalized enrichment scores (NES) were identified for pathways related to ribosomal and mitochondrial processes, while negative NES were identified for pathways involved in immune processes, neuron projection guidance, calcium ion binding, and membrane processes (Figure 1C, Table S2-S4).

**Figure 1.**
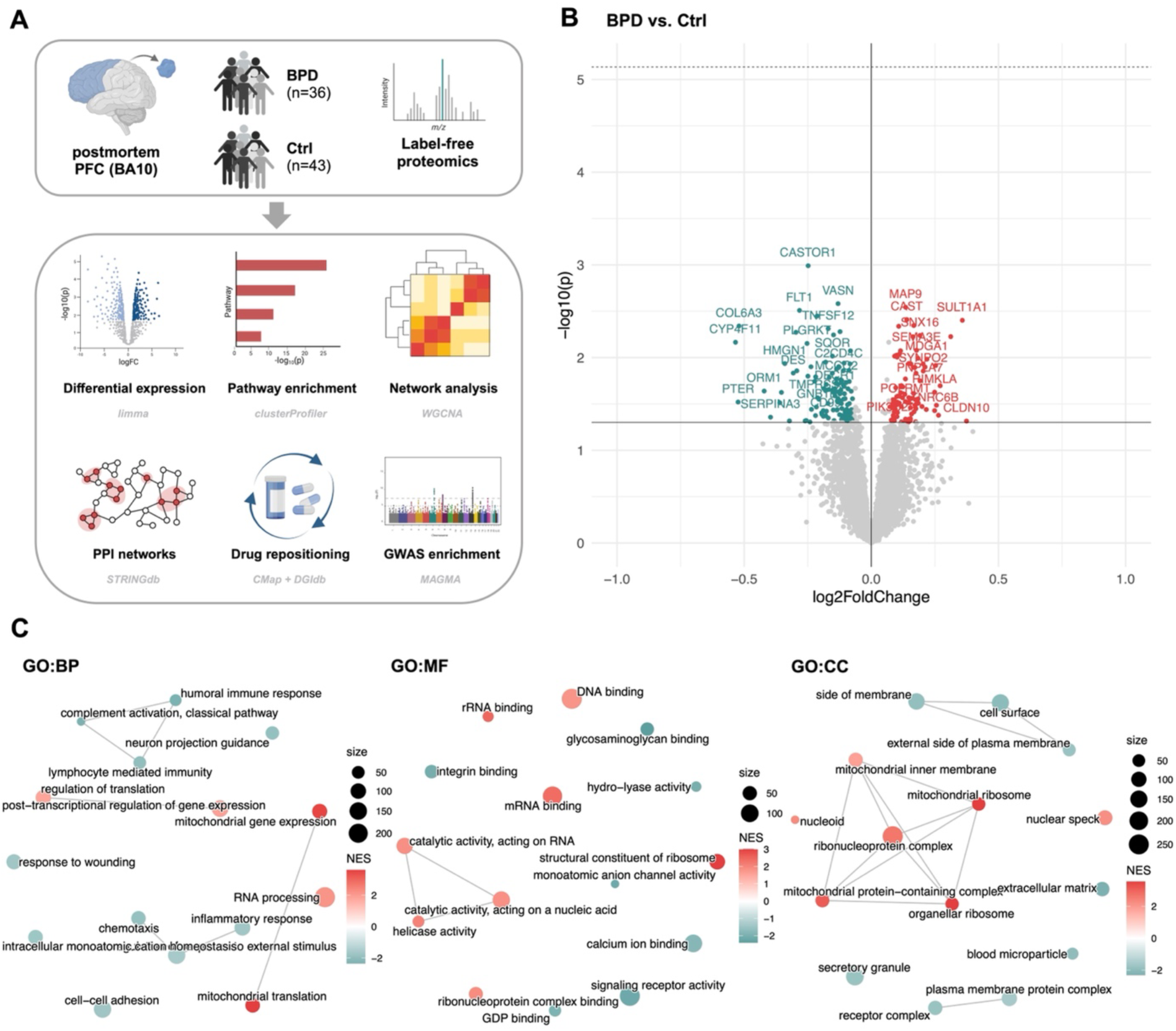
Identification of BPD proteomic signatures by differential protein expression analysis in postmortem PFC. **A** Study and analysis design of the proteomic investigation of BPD in the prefrontal cortex (PFC). **B** Volcano plot of the differential protein expression analysis (BPD vs. Ctrl, n=79) with upregulated proteins highlighted in red and downregulated proteins disoplayed in green. Solid line indicates nominal significance, dashed line 5% FDR significance. **C** Gene set enrichment analysis (GSEA) results separated by Gene Ontology domains Biological Process (BP), Molecular Function (MF), and Cellular Component (CC). Positive and negative GSEA normalized enrichment scores are highlighted in red and green, respectively. BA10 - Brodmann Area 10.

Next, we aimed to identify brain proteomic co-regulation patterns in BPD by investigating co-expression networks in WGCNA (Figure 2A). In our dataset of n=5,580 proteins, where proteins with missing values were excluded to ensure robustness of co-expression networks, we identified a total of 25 protein modules in the PFC (Figure S2A, Table S5). Correlation of module eigengenes with phenotypic variables revealed 7 modules that were significantly associated with BPD (Figure 2B). Analysis of protein overlap across WGCNA co-expression modules and DEPs (p<0.05) suggested modules *blue*, *darkgrey*, *greenyellow*, and *lightyellow* to be enriched for BPD-associated DEPs (Figure 2B+C). Among BPD-associated protein networks, module *lightyellow* was significantly correlated with BPD but not with any other phenotypic variable. We thus further investigated module *lightyellow* by performing a GO enrichment analysis on its 39 defining proteins (Figure 2D, Table S6). Strongest pathway enrichment was found for the GO terms *postsynapse organization* (q=1.69e-02) and *protein localization to endosome* (q=1.59e-02), while other enriched pathways were also related to secretory processes such as *protein targeting to ER* (q=1.69e-02) and *late endosome to lysosome transport* (q=4.91e-02). We further investigated whether WGCNA modules were enriched for BPD-associated DEPs, suggesting dynamic changes in network activity in BPD. We found modules *darkgrey* and *greenyellow* to be significantly enriched for upregulated DEPs, while modules blue and lightyellow were significantly enriched for downregulated DEPs (Figure 2E). Finally, we were interested if proteins of module *lightyellow* are druggable and if so, which drugs are promising to modulate this proteomic co-expression module in BPD (Table S7). Among top DGIdb interaction scores (Figure 2F), we idenfited several drugs for targeting module *lightyellow* proteins NME2 and SRP19. For NME2, antiviral drugs adefovir, tenofovir and zidovudine were identified, while SRP19 is druggable with selective serotonine reuptake inhibitors (SSRI) such as sertraline, paroxetine, and fluoxetine.

**Figure 2.**
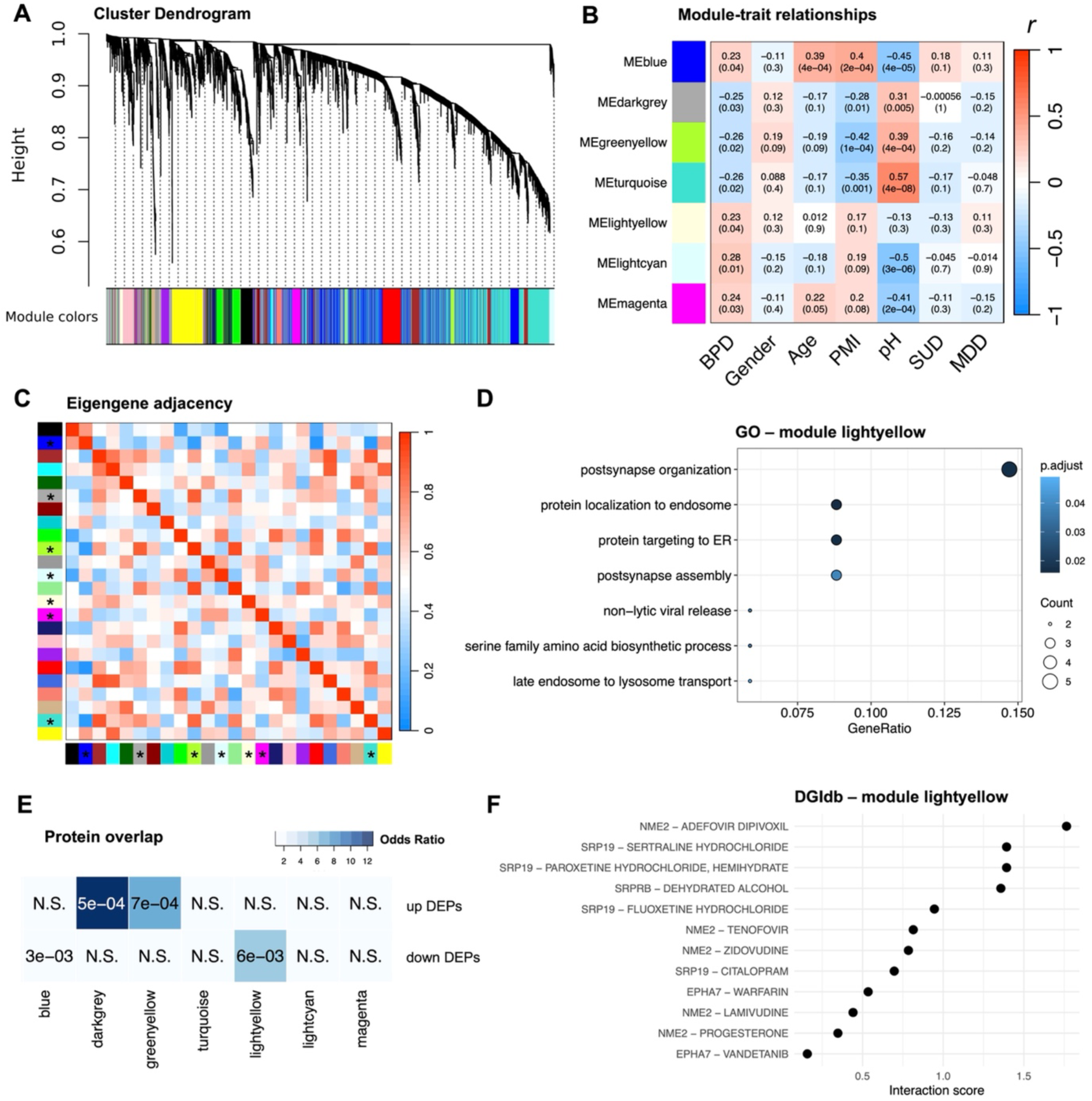
Characterization of WGCNA protein co-expression networks associated with BPD. **A** WGCNA cluster dendrogram for normalized protein intensities of 5,580 proteins in n=79 samples. **B** Correlation of WGCNA protein co-expression module eigengenes with phenotypic variables. Positive Pearson correlation is highlighted in red, negative correlation in blue. Statistical significance of correlation is provided in brackets. BPD: borderline personality disorder, PMI: postmortem interval, SUD: substance use disorder, MDD: major depressive disorder. **C** Eigengene adjacency matrix for the protein co-expression modules in PFC. BPD-associated modules are highlighted with an asterisk. **D** Gene ontology (GO) enrichment analysis of proteins defining BPD-associated module lightyellow. Adjusted p-values highlighted in blue. **E** Protein overlap analysis of WGCNA module proteins and DEPs (nominal p<0.05) from the differential expression analysis. Significant p-values are provided in panels, N.S.: not significant. Odds ratio of overlap highlighted in blue. **F** Results of the druggability analysis in DGIdb for module lightyellow proteins. Results are scored by DGIdb interaction score.

To further investigate the role of protein interactions in BPD, we identified protein-protein interaction (PPI) networks based on BPD-associated DEPs. Using STRINGdb, we performed k-means subclustering on the resulting full PPI network to identify subnetworks related to functional domains (Table S8+S9). The largest PPI subnetworks were related to *mitochondrial translation*, *neurophilin interactions with VEGF and VEGFR*, and *nicotinate and nicotinamide metabolism* (Figure 3A). For the individual PPI subnetworks, we performed a druggability analysis in DGIdb (Figure 3B), suggesting that the subnetwork related to mitochondrial translation is accessible by BAX-targeting drugs, while several tyrosine kinase inhibitors such as pegaptanib, pemigatinib, and tivozanib were suggesting for targeting the subnetwork related to neurophilin interactions with VEGF and VEGFR (Table S10+S11). Finally, for the subnetwork involved in nicotinate and nicotinamide metabolism, we identified immune-modulating drugs such as azathioprine and sulfasalazine (Table S12).

**Figure 3.**
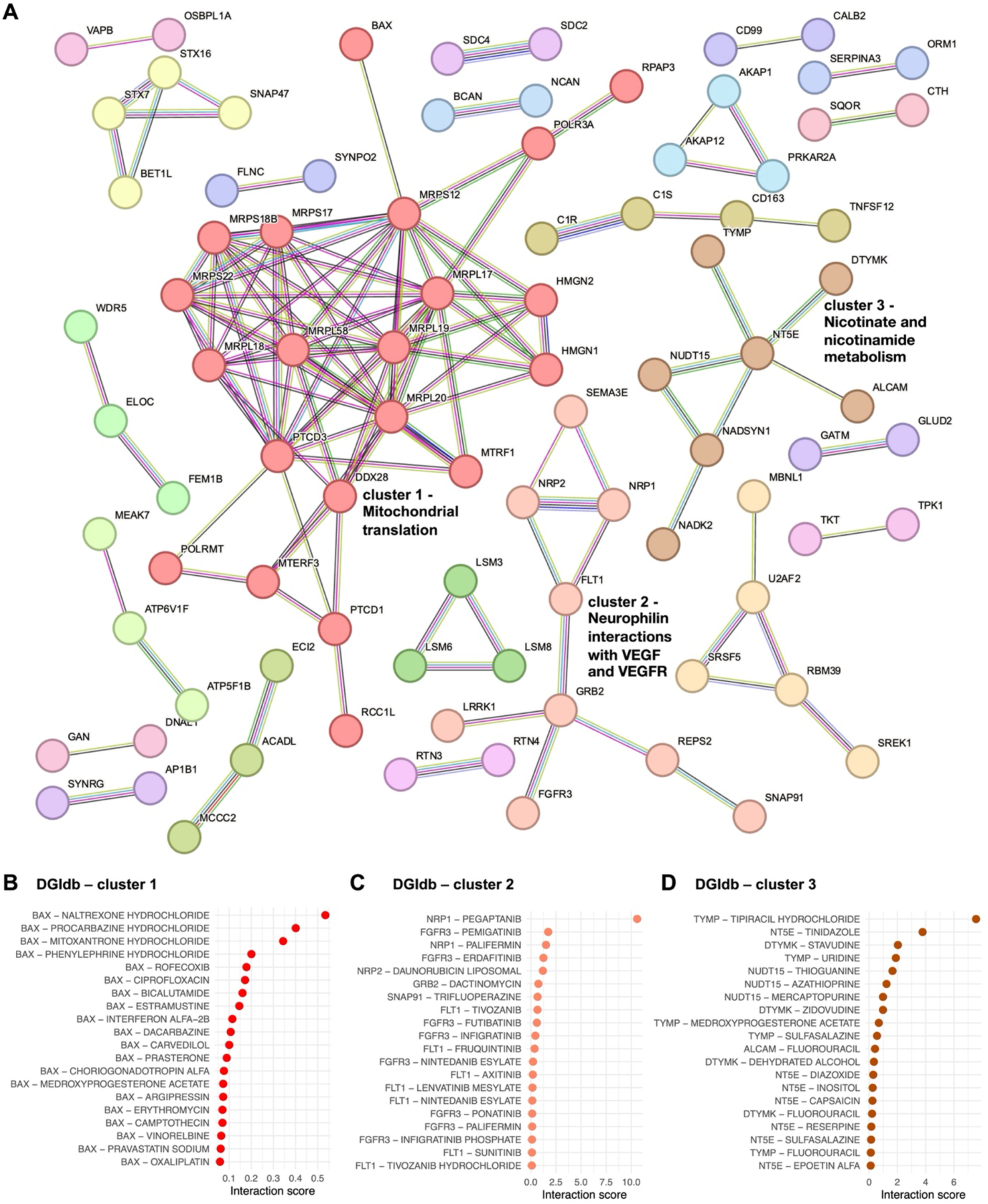
Protein-protein interaction network analysis from differentially expressed proteins in BPD. **A** Protein-protein interaction (PPI) network from STRING for BPD-associated differentially expressed proteins (nominal p<0.05). Subclusters of the interaction network were identified by k-means clustering. The top three largest subclusters are labeled with biological functions. Results of a druggability analysis in DGidb ranked by interaction score (top 20 results) are shown for **B** cluster 1, **C** cluster 2, and **D** cluster 3.

We complemented the druggability analysis in DGIdb by performing a signature-based drug repositioning analysis using connectivity map (CMap). Based on gene expression changes from high-throughput drug screening in the LINCS1000 dataset, we aimed to identify drugs that could reverse the observed protein deregulation patterns in BPD. CMap results suggest VEGFR inhibitors and modulation of the angiopoietin receptor pathway as targets for BPD-associated protein changes (Figure S3A+B), supporting the druggability analysis in PPI network subcluster 2 related to VEGF signaling.

As individuals with BPD frequently present with symptoms of additional psychiatric diagnoses such as PTSD and MDD, we investigated the relationship of brain proteomics signatures of BPD with those from a study investigating the PFC proteome in MDD and PTSD(47). We first overlapped DEPs identified in the individual comparisons BPD vs. Ctrl, PTSD vs. Ctrl, and MDD vs. Ctrl (Table S13). Here, we found two proteins, STX7 and RTN4, that were conserved DEPs across comparisons (Figure 4A). While both proteins were upregulated in BPD, downregulation was observed in PTSD and MDD. In addition, AP1B1 and DDAH2 were shared DEPs across BPD and PTSD, but not MDD, both showing inverse directions of deregulation across phenotypes. Of the 8 DEPs shared between BPD and MDD, 6 (ATP6V1F, MCCC2, ELOC, SGTA, BCAN, and HINT3) were consistently downregulated across disorders. Next to the DEP-focused approach on proteomic overlaps across disorders, we also performed an analysis of convergent and divergent protein deregulation patterns between BPD, MDD, and PTSD using RRHO. Here, instead of investigating individual proteins according to a certain threshold of significance, the analysis is based on gene scoring by both effect size and p-value, thereby providing information on global protein signatures of the phenotype. Using RRHO, we identified a strong convergent overlap of proteomic signatures between BPD and PTSD (Figure 4B), BPD and MDD (Figure 4C), and MDD and PTSD (Figure 4D).

**Figure 4.**
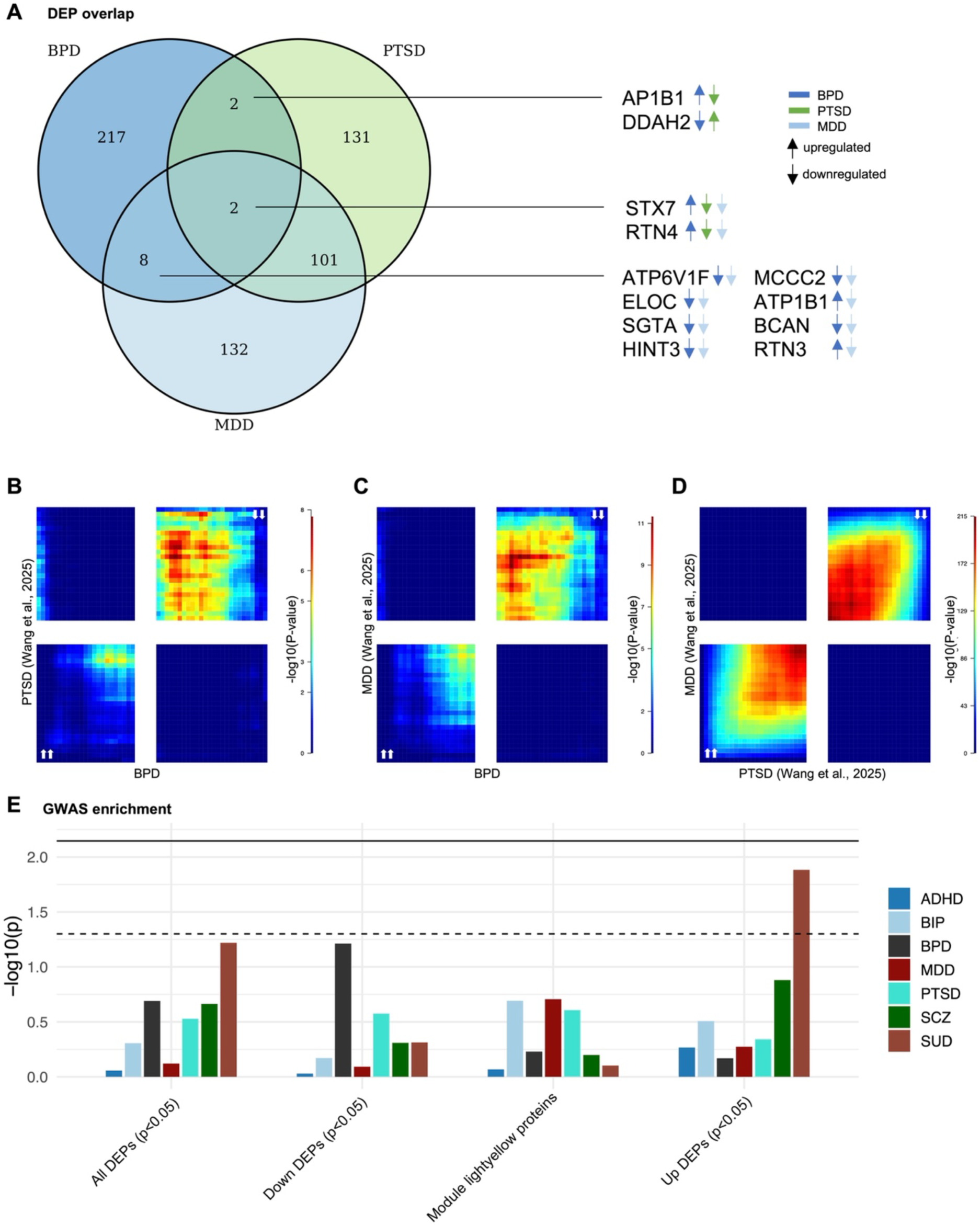
Comparative analysis of BPD proteomic signatures with proteomic and genomic datasets. **A** Venn diagram of BPD-associated differentially expressed proteins (DEPs, nominal p<0.05) overlapped with DEPs from a proteomic study of MDD and PTSD by Wang, Liu (47). Arrows indicate the direction of deregulation in the individual phenotypes. Convergent and divergent protein changes across disorders were investigated using rank-rank hypergeometric overlap analysis (RRHO) for **B** BPD vs. PTSD, **C** BPD vs. MDD, and **D** MDD vs. PTSD comparisons. **E** GWAS enrichment analysis results summarized by enrichment p-values from MAGMA for BPD-associated DEPs (nominal p<0.05) and WGCNA module *lightyellow* proteins. Dashed line indicates nominal significance, solid line indicates significance after Bonerroni-correction for multiple testing.

An important relationship between BPD and other psychiatric diagnoses was also identified by strong genetic correlations for instance with PTSD, MDD, and bipolar disorder (BIP)(11). To integrate protein-level results with genetic data, we performed a GWAS enrichment analysis of BPD DEP signatures and WGCNA module lightyellow proteins in GWAS associations of 7 psychiatric phenotypes including ADHD, BIP, BPD, MDD, PTSD, SCZ, and SUD (Figure 4E, Table S14). Here, we found a significant enrichment of upregulated BPD DEPs in GWAS signals of cross-substance SUD (p=1.31e-02). This association did however not remain statistically significant after Bonferroni correction for multiple testing.

## Discussion

In our brain proteomic study of the prefrontal cortex in BPD, we identified novel associations of protein changes in BPD and identified a convergent overlap of protein signatures with those of frequent co-diagnoses of BPD, such as PTSD and MDD. To our knowledge, it is the first study that describes molecular signatures in the brain of individuals with BPD and therefore adds important evidence to previous association results derived from studies in peripheral blood. We focused our study on the protein level as biological processes such as signaling pathways are largely dependent on the physical interaction of proteins. While studies at the epigenetic and transcript level provide valuable insights into disease processes, they might miss important protein level findings due to complex regulatory processes in biological systems(49). Further, proteins are the major target of pharmaceutical drugs, suggesting a strong translational value of our proteomic analysis.

Differential protein expression analysis identified MAP9 and CASTOR1 as top up- and downregulated DEPs in BPD, respectively. Microtubule-associated protein 9 (MAP9) is an important structural protein important for neuron morphogenesis(50), while Cytosolic Arginine Sensor For MTORC1 Subunit 1 (CASTOR1) is involved in mTOR signaling and was related to neuroinflammation by regulating M1/M2 state switches of microglia(51). In GSEA of differential protein expression signatures, we found positive enrichment scores for ribosomal and mitochondrial processes, which have been related to chronic stress and trauma susceptibility, for instance in PTSD(52) and MDD(53). This finding is supported by results of the PPI network analysis where the strongest connected network was related to mitochondrial translation, therefore suggesting altered mitochondrial protein turnover in the brain of individuals with BPD.

Further, negative enrichment scores were found for pathways involved in immune processes, as well as for neuron projection guidance and calcium ion binding, suggesting that neuroinflammatory and neuroplastic processes are altered in BPD. While these processes have been repeatedly described to be deregulated in psychiatric disorders closely associated with BPD, including PTSD, MDD, and SUDs(54–56), this has not been related to BPD so far. The finding of altered neuroplasticity in BPD is further supported by pathway enrichment in the WGCNA module *lightyellow,* suggesting postsynaptic organization changes in the PFC.

Druggability analysis in WGCNA module *lightyellow* highlighted SSRIs including sertraline, paroxetine, and fluoxetine as pharmaceutical drugs targeting this co-expression network. While SSRIs are often prescribed to BPD patients, they show limited effectiveness for BPD itself, but can be of use if comorbidities such as MDD require treatment(57). A novel and conserved result across druggability analyses in PPI sub-networks and drug repositioning analysis in BPD-associated DEPs were drugs targeting angiogenesis and VEGF signaling pathways. FLT1, the vascular endothelial growth factor receptor 1 (VEGFR1), downregulated in BPD, was among the top 5 significant DEP findings and is druggable by VEGFR inhibitors such as Tivozanib, Lenvatinib, and Fruquintinib. VEGFR inhibitors were also the strongest finding in the perturbagen class analysis of drug repositioning. VEGF signaling is not only involved in angiogenesis, but also has neuroprotective effects and promotes neurogenesis in the brain(58). Elevated plasma levels of VEGF were repeatedly identified in MDD(59, 60) and a study investigating patients with comorbid MDD and BPD confirmed elevated VEGF levels in blood compared to healthy control subjects, but did not find expression differences for VEGF receptors(61). Further, increased VEGF levels were found to be related to antidepressant treatment response(62). While interfering with VEGF signaling in the brain could depict an interesting novel pharamacotherapeutic target, its role in BPD has to be further investigated, as most evidence is derived from MDD and from studies in peripheral blood.

Due to the strong relationship between BPD and other psychiatric disorders such as PTSD and MDD, we investigated the overlap of proteomic signatures across these disorders in the PFC. We identified a strong concordance of brain proteomic signatures across disorders by RRHO. At the level of single proteins, with STX7 and RTN4, we found two proteins that were shared DEPs across BPD, PTSD, and MDD. Syntaxin-7 (STX7) is a member of the SNARE protein family and is involved in vesicular transport and vesicle fusion(63). In rodent hippocampus, Stx7 was related to a presynaptic, rapidly replenishing vesicle pool important for neurotransmission during high-frequency stimuli(64). The ER membrane protein Reticulon 4 (RTN4) is an important regulator of ER morphology and in human cortical neurons was shown to regulate neurite outgrowth and calcium release from the ER(65). While the strong overlap between BPD, PTSD, and MDD has been shown by high rates of co-diagnosis(5) and robust genetic correlations(11), we provide novel insights into convergent protein signatures across these disorders.

Several limitations apply to the present study. First, investigating postmortem brain tissue is associated with the inherent limitation of representing only the endpoint of BPD and it remains unclear if the identified protein signatures reflect causes or consequences of BPD. While there are studies showing stability of protein levels in postmortem brain tissue during the first hours after death(66, 67), there might be proteins that were not detected due to degradation in our samples, and even when adjusted for postmortem interval, non-linear effects of degradation may have influenced the results. Further, due to the limited availability of postmortem brain tissue, our sample size of n=79 remains small, leading to a limited statistical power in differential expression analysis. Finally, BPD depicts a heterogeneous phenotype associated with various psychiatric as well as somatic comorbidities which we aimed to address by aiming for a balanced distribution of comorbidities such as MDD and SUDs across case and control groups and by including frequent them as covariates in our statistical models. Still, strong inter-individual variation in BPD influences the results of association studies, but could be addressed by subgroup analyses in larger samples. Future studies should focus on sex- and ancestry-stratified analyses, which, due to a limited sample size, were not feasible in our cohort. Identifying additional, and even cell type-specific, molecular changes in the brain marks another milestone toward a better understanding of disease processes in BPD. Here, combining multi-omics with single-cell studies depicts an important analytical approach to characterize disease mechnisms and to identify potential targets for novel therapeutic approaches.

## Conclusions

To our knowledge, our study is the first that describes proteomic signatures of BPD in the human brain and thereby contributes to a better understanding of the neurobiology of BPD. It reveals novel protein-level and pathway-level associations, for instance involving immunological, synaptic, and protein secretion pathways. In addition, by suggesting modulation of VEGF signaling, it provides important insights into potential pharmacological treatment options. Finally, our study points towards conserved patterns of protein dysregulation across psychiatric disorders that are frequent comorbidities of BPD.

## Abbreviations

ACE: Adverse Childhood Experience
ADHD: Attention Deficit Hyperactivity Disorder
BA10: Brodmann Area 10
BIP: Bipolar Disorder
BP: Biological Process
BPD: Borderline Personality Disorder
CC: Cellular Component
CMap: Connectivity Map
CoD: Cause of Death
DBB: Douglas Brain Bank
DEP: Differentially Expressed Protein
DIA: Data-independent Acquisition
PFC: Prefrontal Cortex
DSM-IV: Diagnostic and Statistical Manual of Mental Disorders, IV edition
EA: European American
GSEA: Gene Set Enrichment Analysis
GO: Gene Ontology
GWAS: Genome-wide Association Study
Log2FC: Log-2 Fold Change
MAGMA: Multi-marker Analysis of GenoMic Annotation
MDD: Major Depressive Disorder
MF: Molecular Function
NES: Normalized Enrichment Score
PMI: Postmortem Interval
PPI: Protein-Protein Interaction
PTSD: Post-Traumatic Stress Disorder
RRHO: Rank-Rank Hypergeometric Overlap
SCID-II: Structured Clinical Interview for DSM Axis II Personality Disorders
SCZ: Schizophrenia
SSRI: Selective Serotonin Reuptake Inhibitor
SUD: Substance Use Disorder
WGCNA: Weighted Gene Co-Expression Network Analysis

## Declarations

### Ethics approval and consent to participate

Postmortem human brain tissue originated from the Douglas Brain Bank, (Montréal, Canada). Tissue is collected at the brain bank in accordance with established ethical guidelines, including obtaining informed consent from next-of-kin for the use of the tissue for research purposes. The study was approved by the Ethics Committee II of the University of Heidelberg, Medical Faculty Mannheim, Germany, under the register number 2022-566. All methods were carried out in accordance with relevant guidelines and regulations.

### Consent for publication

Not applicable

### Availability of data and materials

The proteomic dataset generated and analysed in the current study is not publicly available due to data privacy reasons, but is available from the corresponding author upon reasonable request.

### Authors’ contributions

Conceptualization, E.Z., D.A., F.Streit., and S.H.W.; Methodology, E.Z., D.A., C.C.W., L.Z., J.F., J.J.S., F. Stein, R.S., F. Streit, and S.H.W.; Resources, N.M., G.T., F. Stein, F. Streit, and S.H.W.; Data curation, E.Z., J.J.S., and F. Stein; Data Analysis, E.Z., and F. Stein; Investigation, E.Z., D.A., C.C.W., L.Z., J.F., J.J.S., F. Stein, R.S., F. Streit, and S.H.W.; Writing – Original Draft, E.Z., D.A., F. Streit, and S.H.W.; Writing – Reviewing & Editing, E.Z., D.A., C.C.W., L.Z., J.F., J.J.S., F. Stein, N.M., G.T., R.S., F. Streit, and S.H.W.; Supervision, J.F., C.C.W., R.S., F. Streit, and S.H.W.; Project Administration, R.S., F. Streit, and S.H.W., Funding Acquisition, R.S., F. Streit, and S.H.W.

## Acknowledgements

Figure 1A was created using BioRender (https://www.biorender.com).

## References

1. Ellison WD, Rosenstein LK, Morgan TA, Zimmerman M. Community and Clinical Epidemiology of Borderline Personality Disorder. Psychiatr Clin North Am. 2018;41(4):561–73.

2. American Psychiatric Association. Diagnostic and statistical manual of mental disorders (5th ed.). 2013.

3. Temes CM, Frankenburg FR, Fitzmaurice GM, Zanarini MC. Deaths by Suicide and Other Causes Among Patients With Borderline Personality Disorder and Personality-Disordered Comparison Subjects Over 24 Years of Prospective Follow-Up. J Clin Psychiatry. 2019;80(1).

4. Álvarez-Tomás I, Ruiz J, Guilera G, Bados A. Long-term clinical and functional course of borderline personality disorder: A meta-analysis of prospective studies. European Psychiatry. 2019;56:75–83.

5. Ramos-Suárez I, Guerrero-Jiménez M, Cervilla JA, Gutiérrez B. Epidemiology of borderline personality disorder in the general population: Prevalence, sociodemographic factors, and comorbidities - A systematic review and meta-analysis. Journal of Affective Disorders. 2026;394:120482.

6. Hall ASM, Musliner KL, Debost J-CP, Vilhjálmsson BJ, Chang Z, D’Onofrio BM, et al. Borderline personality disorder and other psychiatric, somatic, and behavioral conditions: a nationwide family study. Translational Psychiatry. 2026;16(1):257.

7. Hastrup LH, Jennum P, Ibsen R, Kjellberg J, Simonsen E. Borderline personality disorder and the diagnostic co-occurrence of mental health disorders and somatic diseases: A controlled prospective national register-based study. Acta Psychiatr Scand. 2024;149(2):124–32.

8. Schneider F, Erhart M, Hewer W, Loeffler LA, Jacobi F. Mortality and Medical Comorbidity in the Severely Mentally Ill. Dtsch Arztebl Int. 2019;116(23-24):405–11.

9. Bohus M, Stoffers-Winterling J, Sharp C, Krause-Utz A, Schmahl C, Lieb K. Borderline personality disorder. Lancet. 2021;398(10310):1528–40.

10. Skoglund C, Tiger A, Rück C, Petrovic P, Asherson P, Hellner C, et al. Familial risk and heritability of diagnosed borderline personality disorder: a register study of the Swedish population. Mol Psychiatry. 2021;26(3):999–1008.

11. Streit F, Awasthi S, Hall ASM, Braun A, Niarchou M, Marouli E, et al. Genome-wide association analyses of borderline personality disorder identify 11 loci and highlight shared risk with mental and somatic disorders. Nature Genetics. 2026;58(8):1831–44.

12. Porter C, Palmier-Claus J, Branitsky A, Mansell W, Warwick H, Varese F. Childhood adversity and borderline personality disorder: a meta-analysis. Acta Psychiatr Scand. 2020;141(1):6–20.

13. Vonderlin R, Senyüz B, Claus C, Mahalingam S, Schulze A, Koch S, et al. The role of different forms of adverse childhood experiences in borderline personality disorder: results from a network analysis including clinical and non-clinical populations. Borderline Personality Disorder and Emotion Dysregulation. 2026;13(1):20.

14. Drews E, Fertuck EA, Koenig J, Kaess M, Arntz A. Hypothalamic-pituitary-adrenal axis functioning in borderline personality disorder: A meta-analysis. Neurosci Biobehav Rev. 2019;96:316–34.

15. Martín-Blanco A, Ferrer M, Soler J, Salazar J, Vega D, Andión O, et al. Association between methylation of the glucocorticoid receptor gene, childhood maltreatment, and clinical severity in borderline personality disorder. J Psychiatr Res. 2014;57:34–40.

16. Brendel GR, Stern E, Silbersweig DA. Defining the neurocircuitry of borderline personality disorder: functional neuroimaging approaches. Dev Psychopathol. 2005;17(4):1197–206.

17. Schulze L, Schmahl C, Niedtfeld I. Neural Correlates of Disturbed Emotion Processing in Borderline Personality Disorder: A Multimodal Meta-Analysis. Biol Psychiatry. 2016;79(2):97–106.

18. Sicorello M, Schmahl C. Emotion dysregulation in borderline personality disorder: A fronto-limbic imbalance? Curr Opin Psychol. 2021;37:114–20.

19. Paret C, Kluetsch R, Zaehringer J, Ruf M, Demirakca T, Bohus M, et al. Alterations of amygdala-prefrontal connectivity with real-time fMRI neurofeedback in BPD patients. Soc Cogn Affect Neurosci. 2016;11(6):952–60.

20. Dawidowski B, Franczak Ł, Podwalski P, Michalczyk A, Łupkowska-Grygorcewicz A, Piotrowska O, et al. The Epigenetic Landscape of Borderline Personality Disorder: Insights from a Systematic Review. J Clin Med. 2025;14(22).

21. Schmahl C, Arvastson L, Tamm JA, Bohus M, Abdourahman A, Antonijevic I. Gene expression profiles in relation to tension and dissociation in borderline personality disorder. PLoS One. 2013;8(8):e70787.

22. Arranz MJ, Gallego-Fabrega C, Martín-Blanco A, Soler J, Elices M, Dominguez-Clavé E, et al. A genome-wide methylation study reveals X chromosome and childhood trauma methylation alterations associated with borderline personality disorder. Transl Psychiatry. 2021;11(1):5.

23. Prados J, Stenz L, Courtet P, Prada P, Nicastro R, Adouan W, et al. Borderline personality disorder and childhood maltreatment: a genome-wide methylation analysis. Genes Brain Behav. 2015;14(2):177–88.

24. Teschler S, Bartkuhn M, Künzel N, Schmidt C, Kiehl S, Dammann G, et al. Aberrant Methylation of Gene Associated CpG Sites Occurs in Borderline Personality Disorder. PLOS ONE. 2013;8(12):e84180.

25. Sicorello M, Sprenger JC, Störkel LM, Sarg B, Kremser L, Schmahl C, et al. Demonstrating the potential of untargeted hair proteomics for personalized biomarkers in stress-associated disorders. Psychoneuroendocrinology. 2026;184:107684.

26. Sala M, Caverzasi E, Lazzaretti M, Morandotti N, De Vidovich G, Marraffini E, et al. Dorsolateral prefrontal cortex and hippocampus sustain impulsivity and aggressiveness in borderline personality disorder. Journal of Affective Disorders. 2011;131(1):417–21.

27. Soloff PH, Meltzer CC, Becker C, Greer PJ, Kelly TM, Constantine D. Impulsivity and prefrontal hypometabolism in borderline personality disorder. Psychiatry Research: Neuroimaging. 2003;123(3):153–63.

28. Peng K, Steele SC, Becerra L, Borsook D. Brodmann area 10: Collating, integrating and high level processing of nociception and pain. Prog Neurobiol. 2018;161:1–22.

29. Schmahl C, Baumgärtner U. Pain in Borderline Personality Disorder. Mod Trends Pharmacopsychiatry. 2015;30:166–75.

30. R Core Team. R: A Language and Environment for Statistical Computing. R Foundation for Statistical Computing Vienna: R Foundation. 2022.

31. Demichev V, Messner CB, Vernardis SI, Lilley KS, Ralser M. DIA-NN: neural networks and interference correction enable deep proteome coverage in high throughput. Nature Methods. 2020;17(1):41–4.

32. Gatto L, Gibb S, Rainer J. MSnbase, Efficient and Elegant R-Based Processing and Visualization of Raw Mass Spectrometry Data. Journal of Proteome Research. 2021;20(1):1063–9.

33. Ritchie ME, Phipson B, Wu D, Hu Y, Law CW, Shi W, et al. limma powers differential expression analyses for RNA-sequencing and microarray studies. Nucleic Acids Res. 2015;43(7):e47.

34. Yu G, Wang LG, Han Y, He QY. clusterProfiler: an R package for comparing biological themes among gene clusters. Omics. 2012;16(5):284–7.

35. Yu. enrichplot: Visualization of Functional Enrichment Result. R package version 1.24.4, https://yulab-smu.top/biomedical-knowledge-mining-book/. 2024.

36. Langfelder P, Horvath S. WGCNA: an R package for weighted correlation network analysis. BMC Bioinformatics. 2008;9(1):559.

37. Szklarczyk D, Kirsch R, Koutrouli M, Nastou K, Mehryary F, Hachilif R, et al. The STRING database in 2023: protein-protein association networks and functional enrichment analyses for any sequenced genome of interest. Nucleic Acids Res. 2023;51(D1):D638–d46.

38. Subramanian A, Narayan R, Corsello SM, Peck DD, Natoli TE, Lu X, et al. A Next Generation Connectivity Map: L1000 Platform and the First 1,000,000 Profiles. Cell. 2017;171(6):1437–52.e17.

39. Cannon M, Stevenson J, Stahl K, Basu R, Coffman A, Kiwala S, et al. DGIdb 5.0: rebuilding the drug–gene interaction database for precision medicine and drug discovery platforms. Nucleic Acids Research. 2024;52(D1):D1227–D35.

40. de Leeuw CA, Mooij JM, Heskes T, Posthuma D. MAGMA: generalized gene-set analysis of GWAS data. PLoS Comput Biol. 2015;11(4):e1004219.

41. Demontis D, Walters GB, Athanasiadis G, Walters R, Therrien K, Nielsen TT, et al. Genome-wide analyses of ADHD identify 27 risk loci, refine the genetic architecture and implicate several cognitive domains. Nat Genet. 2023;55(2):198–208.

42. O’Connell KS, Koromina M, van der Veen T, Boltz T, David FS, Yang JMK, et al. Genomics yields biological and phenotypic insights into bipolar disorder. Nature. 2025;639(8056):968–75.

43. Trans-ancestry genome-wide study of depression identifies 697 associations implicating cell types and pharmacotherapies. Cell. 2025;188(3):640–52.e9.

44. Nievergelt CM, Maihofer AX, Atkinson EG, Chen CY, Choi KW, Coleman JRI, et al. Genome-wide association analyses identify 95 risk loci and provide insights into the neurobiology of post-traumatic stress disorder. Nat Genet. 2024;56(5):792–808.

45. Trubetskoy V, Pardiñas AF, Qi T, Panagiotaropoulou G, Awasthi S, Bigdeli TB, et al. Mapping genomic loci implicates genes and synaptic biology in schizophrenia. Nature. 2022;604(7906):502–8.

46. Hatoum AS, Colbert SMC, Johnson EC, Huggett SB, Deak JD, Pathak G, et al. Multivariate genome-wide association meta-analysis of over 1 million subjects identifies loci underlying multiple substance use disorders. Nat Ment Health. 2023;1(3):210–23.

47. Wang J, Liu Y, Li H, Nguyen TP, Soto-Vargas JL, Wilson R, et al. A multi-omic approach implicates novel protein dysregulation in post-traumatic stress disorder. Genome Med. 2025;17(1):43.

48. Cahill KM, Huo Z, Tseng GC, Logan RW, Seney ML. Improved identification of concordant and discordant gene expression signatures using an updated rank-rank hypergeometric overlap approach. Sci Rep. 2018;8(1):9588.

49. Zillich E, Artioli A, Rossetti AC, Avetyan D, Belschner H, Frank J, et al. A multi-omics and cell type-specific characterization of the ventral striatum in human cocaine use disorder. Cell Reports. 2025;44(2).

50. Cetin B, Taheri A, Golcuk M, Monroy BY, Fernandes J, Ori-McKenney KM, et al. Structure and mechanism of microtubule stabilization and motor regulation by MAP9. bioRxiv. 2026:2025.11.17.688911.

51. Hu H, Lu X, Huang L, He Y, Liu X, Wang Y, et al. Castor1 overexpression regulates microglia M1/M2 polarization via inhibiting mTOR pathway. Metab Brain Dis. 2023;38(2):699–708.

52. Amorim FE, Rye CS, Milton AL. Mitochondrial dysfunction in PTSD: A mechanism to understand trauma susceptibility? Psychopharmacology. 2026;243(5):965–88.

53. Zhang X, Eladawi MA, Ryan WG, Fan X, Prevoznik S, Devale T, et al. Ribosomal dysregulation: A conserved pathophysiological mechanism in human depression and mouse chronic stress. PNAS Nexus. 2023;2(10):pgad299.

54. Dunn GA, Loftis JM, Sullivan EL. Neuroinflammation in psychiatric disorders: An introductory primer. Pharmacol Biochem Behav. 2020;196:172981.

55. Duman RS, Aghajanian GK, Sanacora G, Krystal JH. Synaptic plasticity and depression: new insights from stress and rapid-acting antidepressants. Nat Med. 2016;22(3):238–49.

56. Lüscher C, Malenka Robert C. Drug-Evoked Synaptic Plasticity in Addiction: From Molecular Changes to Circuit Remodeling. Neuron. 2011;69(4):650–63.

57. Stoffers JM, Lieb K. Pharmacotherapy for Borderline Personality Disorder—Current Evidence and Recent Trends. Current Psychiatry Reports. 2014;17(1):534.

58. Jin K, Zhu Y, Sun Y, Mao XO, Xie L, Greenberg DA. Vascular endothelial growth factor (VEGF) stimulates neurogenesis in vitro and in vivo. Proc Natl Acad Sci U S A. 2002;99(18):11946–50.

59. Song X, Xiao S, Wang Z, Miao W. The role of vascular endothelial growth factor (VEGF) in major depressive disorder: an update meta-analysis. BMC Psychiatry. 2026.

60. Carvalho AF, Köhler CA, McIntyre RS, Knöchel C, Brunoni AR, Thase ME, et al. Peripheral vascular endothelial growth factor as a novel depression biomarker: A meta- analysis. Psychoneuroendocrinology. 2015;62:18–26.

61. Kahl KG, Bens S, Ziegler K, Rudolf S, Kordon A, Dibbelt L, et al. Angiogenic factors in patients with current major depressive disorder comorbid with borderline personality disorder. Psychoneuroendocrinology. 2009;34(3):353–7.

62. Clark-Raymond A, Meresh E, Hoppensteadt D, Fareed J, Sinacore J, Garlenski B, et al. Vascular endothelial growth factor: Potential predictor of treatment response in major depression. World J Biol Psychiatry. 2017;18(8):575–85.

63. Mullock BM, Smith CW, Ihrke G, Bright NA, Lindsay M, Parkinson EJ, et al. Syntaxin 7 is localized to late endosome compartments, associates with Vamp 8, and Is required for late endosome-lysosome fusion. Mol Biol Cell. 2000;11(9):3137–53.

64. Mori Y, Takenaka K-i, Fukazawa Y, Takamori S. The endosomal Q-SNARE, Syntaxin 7, defines a rapidly replenishing synaptic vesicle recycling pool in hippocampal neurons. Communications Biology. 2021;4(1):981.

65. Konno T, Parutto P, Crapart CC, Davì V, Bailey DMD, Awadelkareem MA, et al. Endoplasmic reticulum morphology regulation by RTN4 modulates neuronal regeneration by curbing luminal transport. Cell Reports. 2024;43(7).

66. ElHajj Z, Cachot A, Müller T, Riederer IM, Riederer BM. Effects of postmortem delays on protein composition and oxidation. Brain Res Bull. 2016;121:98–104.

67. Machaalani R, Gozal E, Berger F, Waters KA, Dematteis M. Effects of post-mortem intervals on regional brain protein profiles in rats using SELDI-TOF-MS analysis. Neurochemistry International. 2010;57(6):655–61.

