## Supplementary Figures for "Proteomic profiling of postmortem prefrontal cortex identifies molecular signatures of borderline personality disorder"

**A**

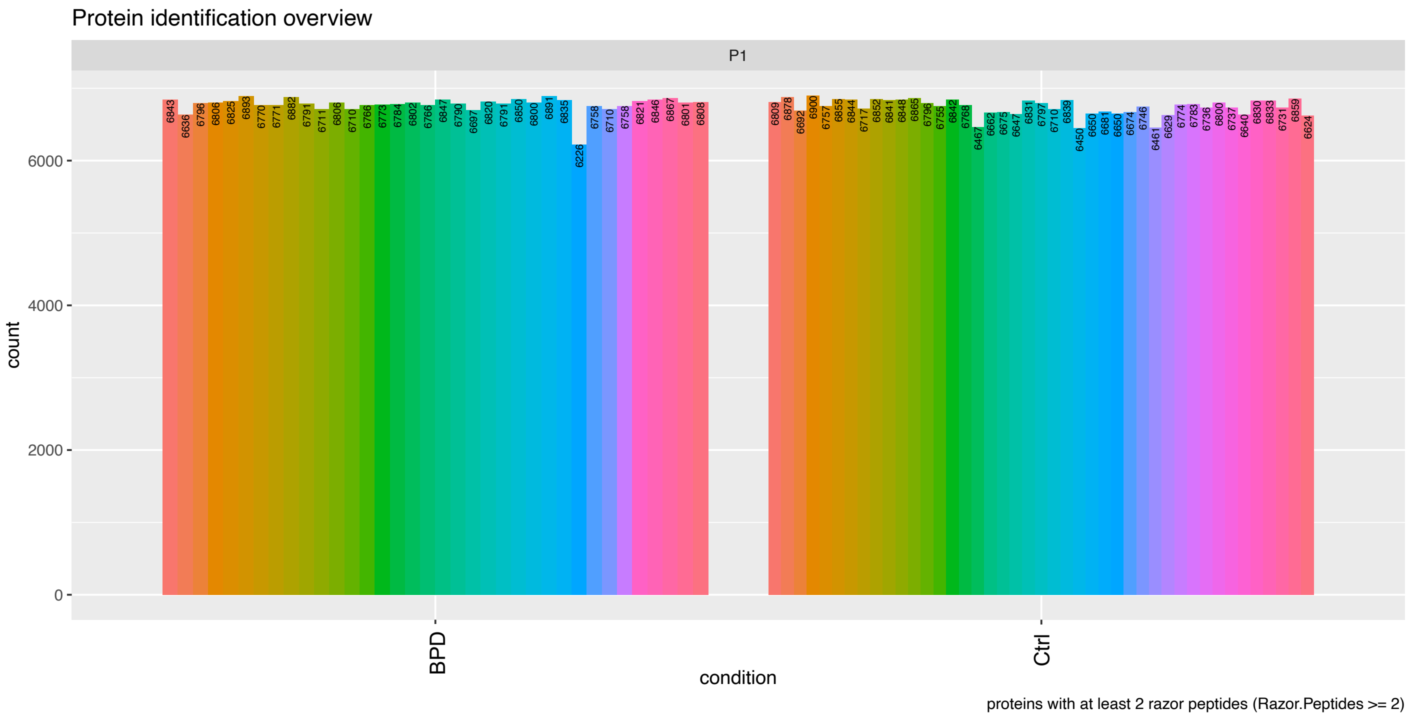


**Supplementary Figure 1 – Number of identified proteins across samples**

**A** For each of the n=79 postmortem brain samples of the prefrontal cortex, the total number of identified proteins per sample is displayed. BPD: borderline personality disorder samples, Ctrl: control samples.

A

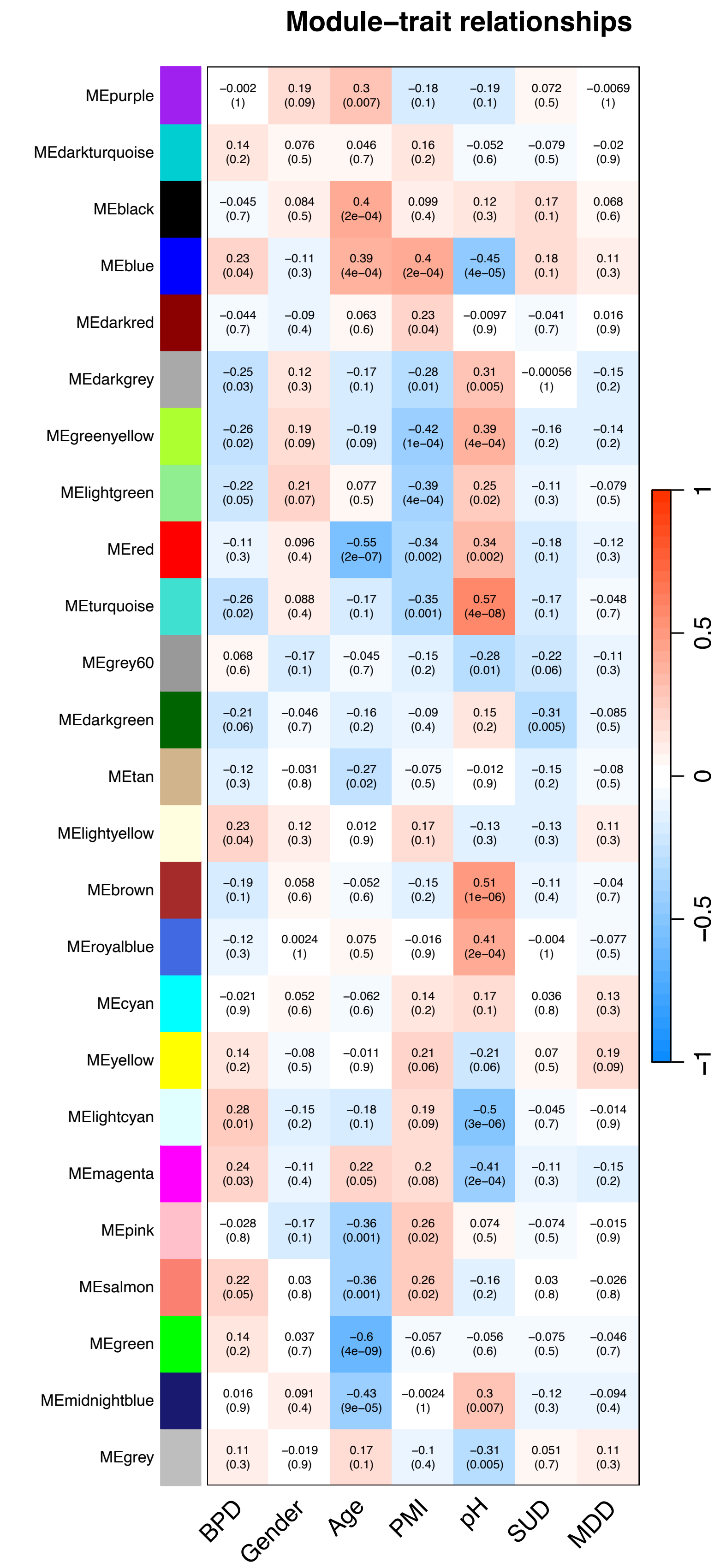


**Supplementary Figure 2 – Module-trait correlation summary for WGCNA protein modules**
**A** Summary of Pearson correlation between module eigengenes of the 25 protein co-expression modules from WGCNA and phenotypic variables. Red color indicates positive, blue color negative Pearson correlation coefficients. Statistical significance of correlation (p-value) is displayed in brackets. BPD: borderline personality disorder, PMI: postmortem interval, pH: brain pH valie, SUD: substance use disorder diagnosis, MDD: major depressive disorder diagnosis.

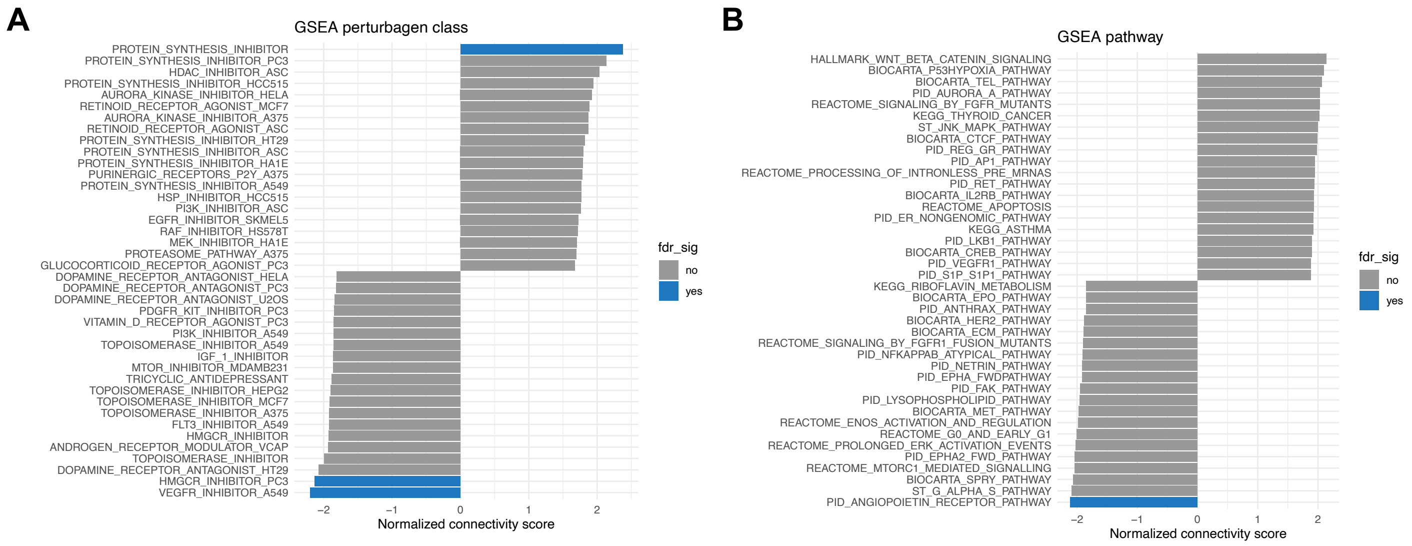


**Supplementary Figure 3 – Results of the drug repositioning analysis using CMap**
Signature-based drug repositioning analysis results were derived from CMap (https://clue.io). Gene set enrichment analysis (GSEA) results for **A** perturbagen class and **B** pathways are displayed and colored by FDR significance of enrichment (fdr_sig, blue: yes, grey: no).
